# Excess mortality and years of life lost from 2020 to 2023 in France: a cohort study of the overall impact of the COVID-19 pandemic on mortality

**DOI:** 10.1101/2023.12.13.23299903

**Authors:** Paul Moulaire, Gilles Hejblum, Nathanaël Lapidus, the COVID HOSP working group

**Author notes:** **Correspondence:** Paul Moulaire, Inserm / Sorbonne Université UMR S 1136, 27 rue Chaligny, 75012 Paris, France.

## Abstract

**Introduction:** Excess mortality has been frequently used worldwide for summarizing the COVID-19 pandemic-related burden. Estimates for France for the years 2020 to 2022 vary substantially from one report to another, and the year 2023 is poorly documented. The present study assessed the level of excess mortality that occurred in France between 2020 and 2023 together with the corresponding years of life lost (YLL), in order to provide a reliable, detailed, and comprehensive description of the overall impact of the pandemic.

**Method:** This open cohort study of the whole French population analyzed the 8,451,372 death occurrences reported for years 2010 to 2023. A Poisson regression model was trained with years 2010 to 2019 for determining the age-and sex-specific evolution trends of mortality before the pandemic period. These trends were then used for estimating the excess mortality during the pandemic period (years 2020 to 2023). The life expectancies of the persons in excess deaths were used for estimating the corresponding years of life lost (YLL).

**Results:** From 2020 to 2023, the number of excess deaths (mean [95% CI] (percentage of change versus expected mortality)) was respectively 49,541 [48,467; 50,616] (+8,0%), 42,667 [41,410; 43,909] (+6.9%), 53,129 [51,696; 54,551] (+8.5%), and 17,355 [15,760; 18,917] (+2.8%). Corresponding YLL were 512,753 [496,029; 529,633], 583,580 [564,137; 602,747], 663,588 [641,863; 685,723], and 312,133 [288,051; 335,929]. Individuals younger than 60 years old accounted for 17% of the YLL in 2020, 26% in 2021, 32% in 2022 and 50% in 2023. Males were more affected than females by both excess mortality and YLL.

**Conclusion:** This study highlights the long-lasting impact of the pandemic on mortality in France, with four consecutive years of excess mortality and a growing impact on people under 60, particularly males, suggesting lasting and profound disruption to the healthcare system.

**Key Messages:** 

**What is already known on this topic:** Different trends and magnitudes of excess mortality were reported in France for years 2020 to 2022, and estimations of years of life lost, which characterize the remaining life expectancy of people suffering excess mortality, are only available for 2020 and 2021. There is no exhaustive toll covering all years of the COVID-19 pandemic period (2020– 2023).

**What this study adds:** Excess mortality peaked in 2022 and remained substantial in 2023 for the fourth year in a row while corresponding years of life lost rose steadily from 2020 to 2022 and remained at a worrying level in 2023.

**How this study might affect research, practice or policy:** This study raises concerns about a potential indirect and long-lasting impact of the COVID-19 pandemic on mortality in France, particularly in males under 60 years old.

## Introduction

The COVID-19 pandemic quickly spread worldwide in 2020. According to the World Health Organization (WHO), the pandemic cumulative direct death toll on December 31, 2023, was 7,015,947 deaths [1]. Excess mortality, defined as the difference between observed and expected mortality, constitutes an attractive feature for summarizing the major impact of the pandemic, for at least two reasons. First, estimating excess mortality only requires all-cause mortality data during a reference period and during the pandemic period of interest. Second, excess mortality is a straightforward estimate of the global burden, including both virus-related direct deaths and indirect deaths related to all perturbations that simultaneously occurred during the pandemic period. Thanks to its relevancy, excess mortality has rapidly become an essential indicator for assessing the impact of the pandemic [2]. As detailed by Vanella et al. in their review article, the concept of excess mortality initially emerged in 1930 and called a particular renewed attention during the COVID-19 pandemic [3].

Numerous studies already estimated excess mortality in France between 2020 and 2021 with heterogeneous results. This heterogeneity is inducted by different assumptions and methodological choices [4–6], as illustrated in the study of Levitt et al. [7], which details the differences between final estimates issued from four major contributions (the COVID-19 Excess Mortality Collaborators [8], Karlinsky and Kobak [9], The Economist team [10], WHO [11]): in France, estimates vary almost three-fold, from 57,767 to 155,000 excess deaths in 2020 and 2021. Considering year 2022, all sources agreed about the maintenance of some excess mortality, but with relative estimates contrasting substantially: The French National Institute for Statistic and Economic Studies (Insee) estimated that excess mortality in 2022 was greater than its corresponding estimates for the two previous pandemic years [12], while in contrast, Eurostat and the European Mortality Monitoring Project estimated that it was lower than in 2020 [13,14]. All these results demonstrate a high variability of estimates depending on methodological choices and raise questions about the real level of excess mortality in France and its evolution during the pandemic period. Concerning 2023, while WHO declared that COVID-19 no longer constituted a public health emergency of international concern (PHEIC) [15], no official estimation of excess mortality in France is published to our knowledge. Furthermore, while excess mortality is a good summary indicator, it provides insufficient information to fully quantify the burden linked to the pandemic since, as underlined by Ferenci [16], raw death counts do not acknowledge the age of the deceased persons. A few studies already investigated the numbers of years of life lost (YLL) related to the pandemic[17,18], but little is known after 2020.

To sum-up, there is a wide range of estimates of excess mortality in France from 2020 to 2022, no official figures for 2023 and a lack of detail on the age and sex structure of people affected after 2020. To address these issues, the study reported here had three objectives.

The first objective was to accurately estimate excess mortality, taking into account changes in population structure over time and natural trends in mortality, in order to shed light on the exact level of excess mortality in France between 2020 and 2022. The second objective was to estimate the level of excess mortality in 2023, in order to assess whether the situation is returning to normal after three consecutive years of increased mortality. The third objective was to estimate the YLL between 2020 and 2023, taking into account the age and sex of those in excess mortality in the balance sheet. By simultaneously addressing these three objectives, the overall aim of the study was to draw up a complete assessment of the pandemic burden in France, from the year of the emergence of COVID-19 in 2020 to the year WHO declared the end of the PHEIC.

## Materials and Methods

### Data source and availability

All data used in the study are available in open access from the French National Institute for Statistic and Economic Studies (Insee), including daily mortality by age and sex [19], population structure [20] and mortality tables [21].

### Participants

The present open cohort study of the French population between 2010 and 2023 is reported according to STROBE guidelines [22]. Throughout the study, the term “sex” refers to the sex assigned at birth. In accordance to the Sex and Gender Equity in Research (SAGER) guidelines [23] sex was precisely taken into account in the model, and the results were detailed by sex. In the above-mentioned databases, the population size by age and sex at a given year consider the situation on January 1.

### Data processing

As the age structure of the French population is given on January 1 of each year, the age of a deceased person considered in the study was that on January 1 of the year of death, and mortality data were aggregated by year. This annual aggregation allowed to smooth the periodicity of the mortality, avoided the short-term harvesting effect (for instance, counting as excess deaths the persons who died prematurely in April 2020 but who would have died within 2020), and provided a toll easier to interpret than monthly or weekly tolls. Persons aged 99 years or more were handled in a single age category and individuals who died outside France were not included in the study. Given that in France the legal deadline for transmitting death certificates to INSEE is one week [24], and that the extraction of mortality data dates from June 2024, the number of missing data between 2010 and 2023 should be minimal.

### Outcomes

For each sex i, age j and year k, expected mortality 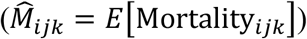 was estimated with a Poisson regression model described below. The numbers of excess deaths 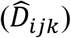 were estimated for each stratum by subtracting the expected mortality from the observed one.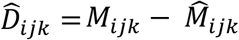. For a given year, the total burden is the sum of the age- and sex-specific excess deaths, e.g.

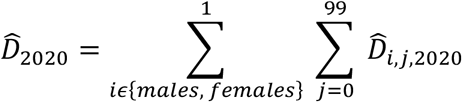

YLL were estimated by multiplying each estimated excess deaths by the corresponding life expectancy (*E*_*ijk*_), based on the mortality table of 2019 [21].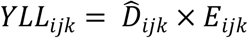. The estimate of the total burden for a given year is the sum of the age- and sex-specific YLL.

### Statistical model and selection process

A Poisson regression model was used to estimate expected mortality in the pandemic period (years 2020 to 2023), with years 2010-2019 handled as the reference period. Such models have already been used to estimate expected level of mortality in order to estimate excess cause specific [25] or all causes excess mortality [26,27]. A ten year-duration of a reference period have also been used in previous studies [8,12].

Age (as a year-specific categorical variable), sex, and year (as a continuous variable) were used as predictors to estimate mortality. Different interactions of these variables were tested, and AIC was used as selection criteria to optimize goodness-of-fit and avoid overfitting. Details of the model selection are given in the Supplementary Material (See Supplementary Text 1 and Supplementary Table 1). The Model used was:

**Table 1.** Excess mortality and corresponding years of life lost by sex, France, 2020−2023.

| Year | 2020 | 2021 | 2022 | 2023 |
| --- | --- | --- | --- | --- |
| **Excess mortality\*** | 49,541 [48,467; 50,616] (+8,0%) | 42,667 [41,410; 43,909] (+6.9%) | 53,129 [51,696; 54,551] (+8.5%) | 17,355 [15,760; 18,917] (+2.8%) |
| Males | 28,925 [28,164; 29,677] (+9.4%) | 28,070 [27,209; 28,839] (+9.2%) | 29,572 [28,578; 30,566] (+9.6%) | 11,623 [10,512; 12,741] (+3.8%) |
| Females | 20,616 [19,841; 21,402] (+6.6%) | 14,597 [13,706; 15,465] (+4.7%) | 23,557 [22,545; 24,575] (+7.5%) | 5,732 [4,589; 6,847] (+1.8%) |
| **Years of life lost†** | 512,753 [496,029; 529,633] | 583,580 [564,137; 602,747] | 663,588 [641,863; 685,723] | 312,133 [288,051; 335,929] |
| Males | 319,011 [306,331; 331,834] | 389,059 [374,698; 403,427] | 398,672 [382,659; 414,739] | 213,911 [196,033; 231,583] |
| Females | 193,742 [182,580; 205,027] | 194,521 [181,692; 207,371] | 264,916 [250,489; 279,666] | 98,222 [81,989; 114,361] |

\*Excess mortality [95% confidence interval] (percentage of increase compared to expected mortality)
†Years of life lost [95% confidence interval]

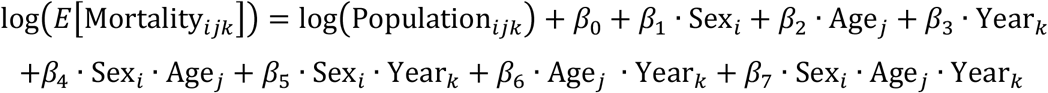

This model allowed to accurately estimate age- and sex-specific expected number of deaths, excess mortality and corresponding years of life lost, considering mortality trends and changes in population structure over time.

### Software, packages and estimation of confidence intervals

All analyses were performed with statistical software R 4.0.2 (R Foundation for Statistical Computing, Vienna, Austria), package ggplot2 was used to generate figures, package stats was used for the general linear model, and 95% confidence intervals (CIs) were estimated over 10,000 bootstrap replications. The use of the viridis color palette in the graphs was based on its perceptual uniformity and colorblind-friendliness to ensure accurate data interpretation and broad accessibility.

## Results

### Population under study

Between 2010 and 2023, the French population grew from 64.6 million to 68.1 million inhabitants, and the corresponding observations included in the study totalized 931,741,878 person-years (see study flow diagram in the Supplementary Figure 1). Over the analysis period, 8,536,511 deaths occurred, of which less than 1% were not considered because they took place abroad. The study was therefore based on 8,451,372 deaths to estimate excess mortality and YLL. A graph plotting the raw mortality events occurring during the study period already documents substantially some mortality patterns in France over time: Figure 1 shows the presence of periodical patterns of mortality, with higher levels usually observed during winters, reflecting, at least in part, the impact of seasonal-circulating viruses such as influenza. Moreover, the figure also evidences the impact of population structure on crude mortality over time. The two dark diagonal stripes (in 2010: one beginning between 64 and 69 years old, and one between 91 and 94 years old) showing lower mortality levels corresponds to two particular cohorts, persons born in 1939–1945 (second world war) and 1914–1918 (first world war). During these two periods, natality was low, inherently resulting in cohorts with smaller sample sizes and a corresponding lower crude mortality over time. Impact of the COVID-19 pandemic is also flagrant in 2020 with, for instance, the especially high mortality in April, followed by a succession of disturbances, until 2023.

**Figure 1:**
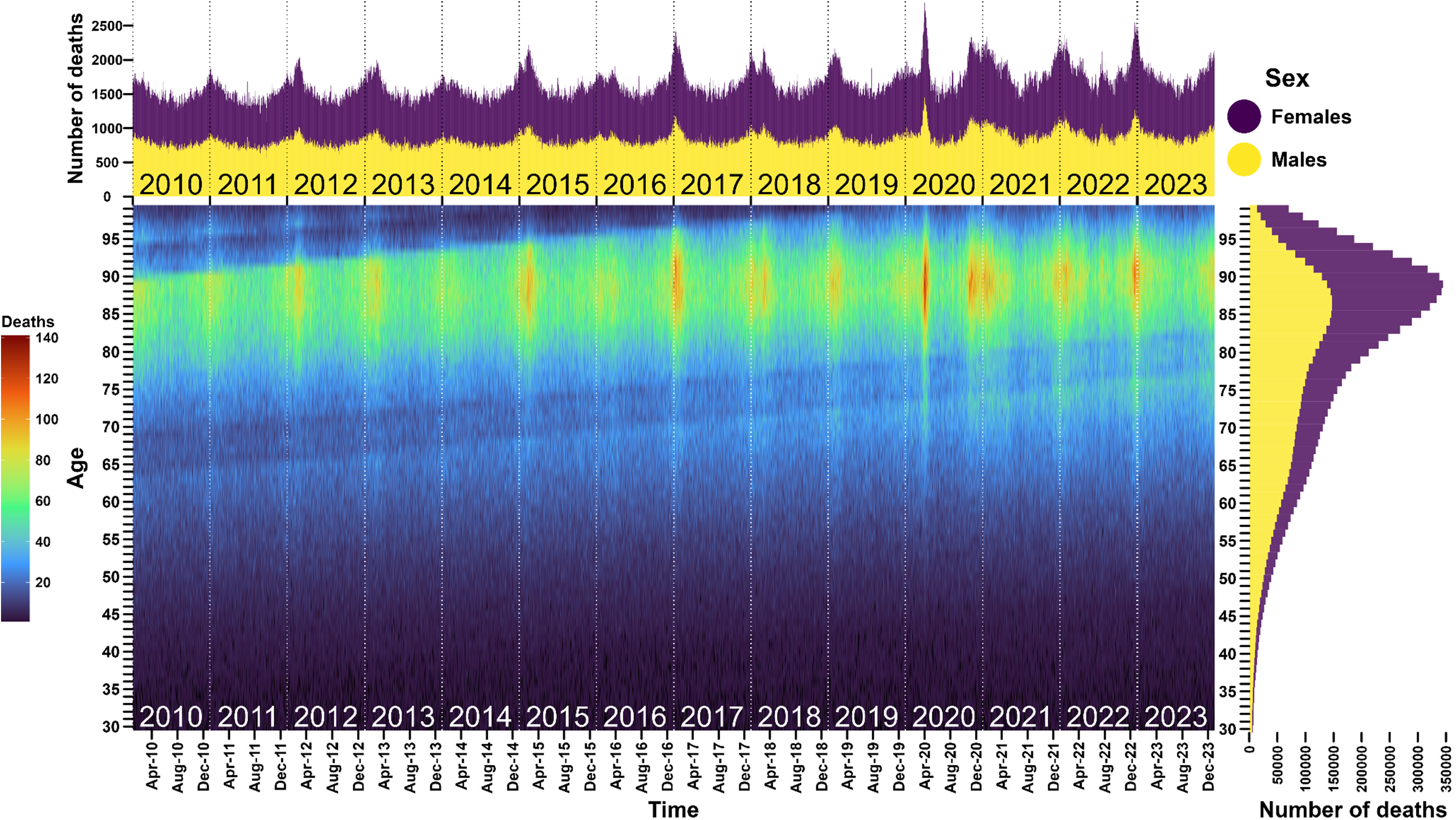
Daily mortality by age, and repartition by sex, France, 2015–2023 Top panel, daily mortality in France from 2015 to 2023 with the repartition by sex; right panel, cumulative number of deaths by exact age from 30 to 99 years old with the repartition by sex; bottom panel, daily number of deaths according to exact age, for instance, in April 2020 the colour gradient is red for people between 85 and 90 years old corresponding to a daily number of deaths around 140 and this mortality peak is also visible in the top panel both for males and females.

### Excess mortality

A total of 162,692 [159,951; 165,335] excess deaths were estimated from 2020 to 2023 with 60% occurring in males. The estimated number of excess deaths (mean [95% CI]) was 49,541 [48,467; 50,616] in 2020, decreased to 42,667 [41,410; 43,909] in 2021, reached a maximum value in 2022 with 53,129 [51,696; 54,551] deaths, before dropping to 17,355 [15,760; 18,917] deaths in 2023 (Table 1). As compared to the expected numbers of deaths, the aforementioned numbers of excess deaths in years 2020–2023, corresponded to increases of 8,0%, 6.9%, 8.5%, and 2.9%, respectively. Figure 2 and upper part of Table 1 also provides insights on the variability of excess mortality over years 2020–2023, and according to sex. Variation of excess mortality between 2020 and 2022 was driven by females: excess mortality in males was almost as the same level during this period, while that in females dropped in 2021 before peaking in 2022. In 2023, excess mortality mainly affected males, with more than twice as many excess deaths as females. As shown in Figure 2, excess mortality principally affected individuals older than 60 years old, but a signal is also visible in males under 60 years old. As detailed in the Supplementary Table 2, individuals in excess mortality were younger in 2020 and 2022 (median ages of 83 and 81 years old, respectively), as compared to 2021 and 2023 (median ages of 78 and 76 years old).

**Figure 2:**
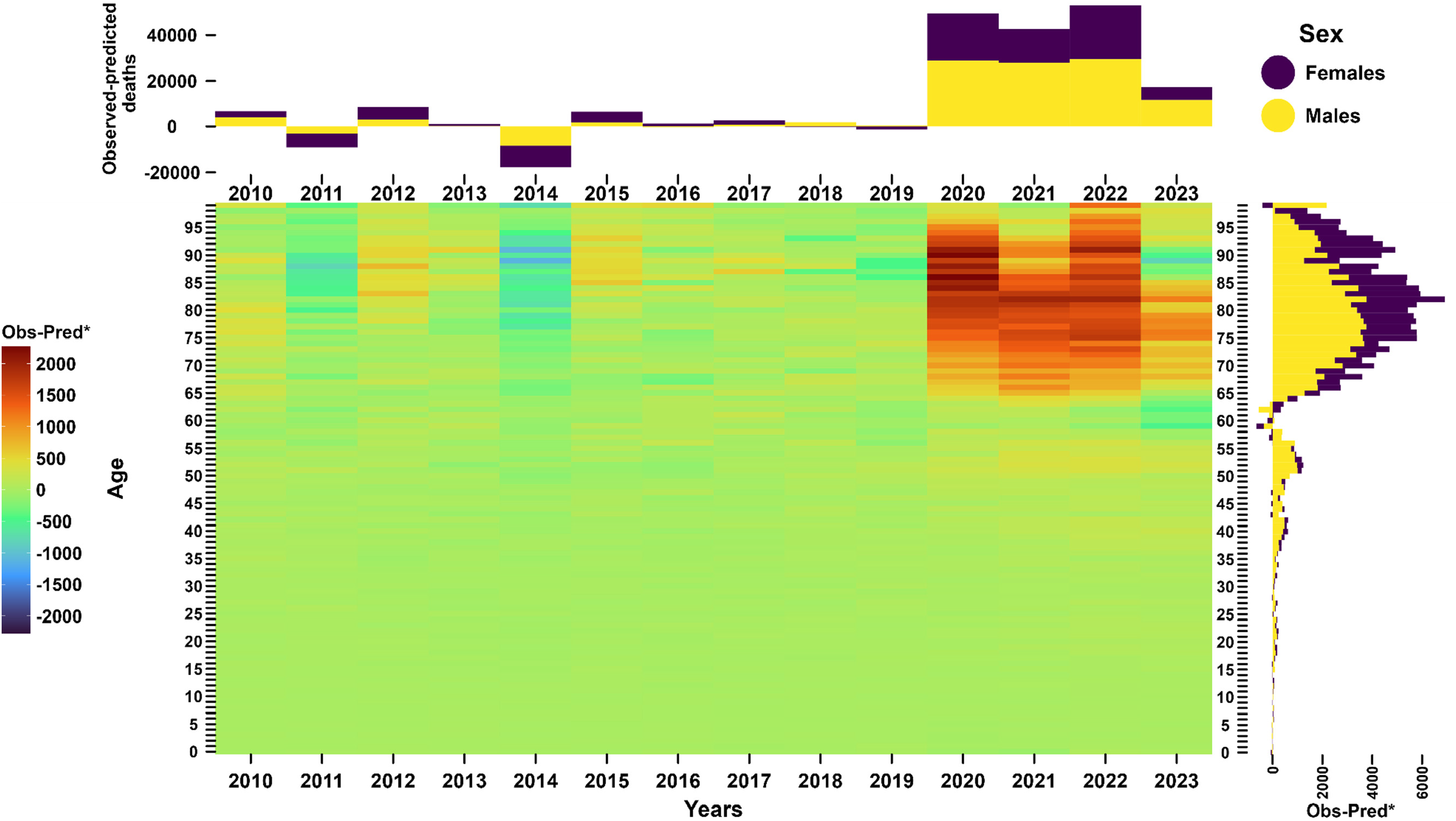
Differences between observed and predicted deaths in France, 2015–2023. *Observed minus predicted number of deaths Top panel, annual cumulative difference between observed and predicted deaths, training period 2010–2019, with the repartition by sex; right panel, cumulative difference between observed and predicted deaths by exact age and according to sex from 2020 to 2023; main panel, annual difference between observed and predicted deaths by exact age, for instance, in 2020, the colour gradient is dark red for people aged 90 years old, corresponding to a difference between observed and predicted number of deaths at about 2000; this difference was considered as excess mortality as the baseline is estimated in pre-pandemic period and corresponds to mortality level in the absence of pandemic.

### Years of life lost

From 2020 to 2023, the estimated total number of YLL related to excess deaths was 2,072,054 [2,030,339; 2,113,188] among which 64% concerned males. As detailed in the lower part of Table 1, the overall level steadily rose from 512,753 [496,029; 529,633] in 2020 to 663,588 [641,863; 685,723] in 2022, before dropping to 312,133 [288,051; 335,929] in 2023. Despite males have a lower life expectancy than females at the same age, the estimated contribution of males to excess death-related YLL during years 2020 to 2023 was always greater than that of females, particularly in 2023. Individuals in excess mortality were younger in 2021 and 2023 than in 2020 and 2022, resulting in a greater mean number of YLL per individual in 2021 (11.2 years) and 2023 (13.1 years) than in 2020 (8.5 years) and 2022 (9.8 years). The contribution of individuals younger than 60 years old to the annual toll of excess mortality-related YLL dramatically rose over time, from 17% in 2020 to 50% in 2023, mainly driven by males from 20 to 60 years old (Figure 3, Supplementary Table 2, and Supplementary Figure 2).

**Figure 3:**
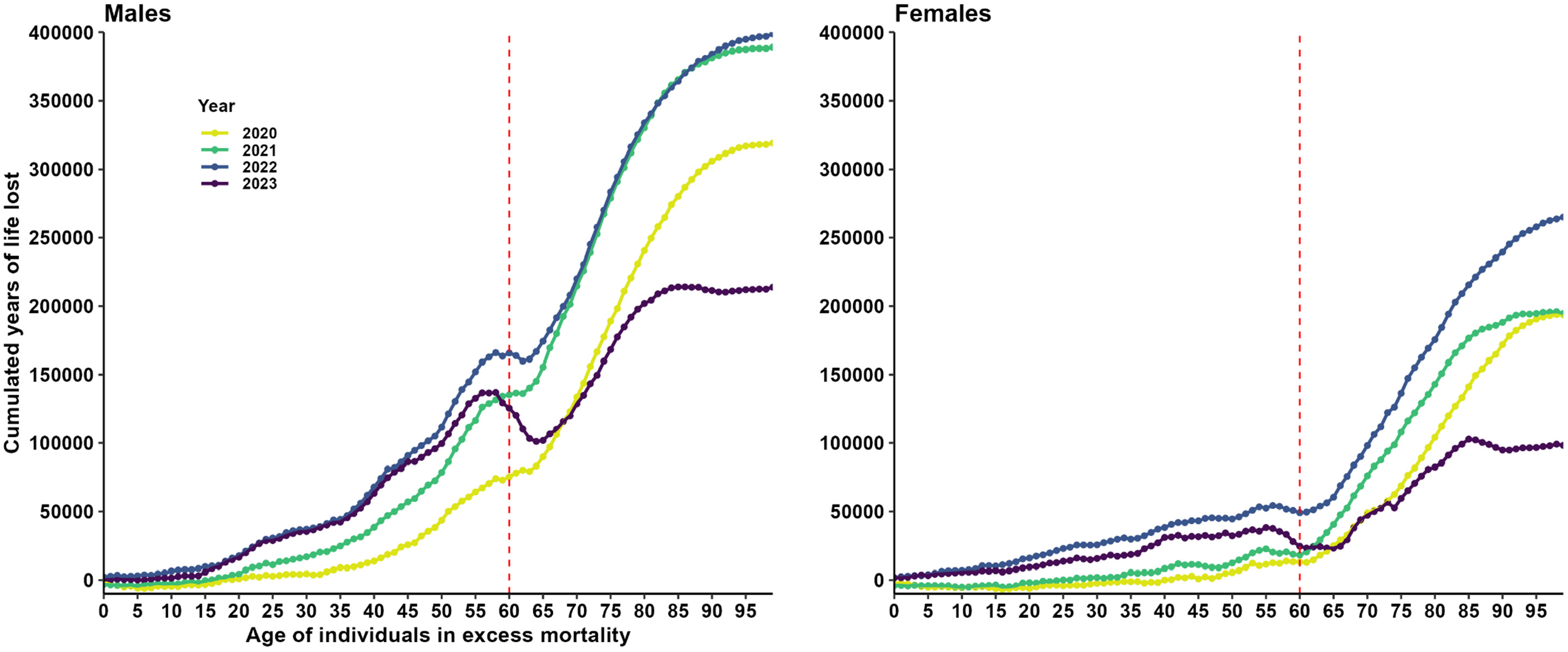
Cumulated number of years of life lost per age of individuals in excess mortality. For each year (color) and each sex (panel) the points represent the cumulated number of years of life lost for individuals in excess mortality from 0 year old to the corresponding age. For instance, in 2020 (yellow line) males (left panel) younger than 60 years old (red dotted line) cumulated 73,000 years of life lost, around 23% of the 319,000 YLL cumulated by males in excess mortality in 2020. Cumulated number of YLL is decreasing when observed number of deaths in a particular stratum is below the expected one (negative excess mortality) as for instance in 2023 for individuals aged 60 years old. Contribution of each age to YLL is detailed in Supplementary Figure 2.

## Discussion

### Main findings

In the present study, excess mortality and corresponding YLL from 2020 to 2023 in France were accurately assessed by considering in the modelling approach both the evolving trend of mortality over the ten last years preceding pandemic, and the changes of population structure over time. In our study, a total of 92,208 excess deaths were estimated for years 2020 to 2021, between the 81,849 excess deaths estimate by the WHO and the 96,831 excess deaths estimated by Levitt [7]. Considering year 2022, the present study estimate of 53,129 excess deaths is close to the 53,800 excess deaths estimated by the Insee [12] and confirms that the level of excess mortality was higher in 2022 than in 2020 and 2021. The present study estimated the occurrence of 17,355 excess deaths in 2023, much less than the estimates for the three previous years. Despite the decline in the number of excess deaths estimated in 2021, the number of YLL constantly increased from 2020 to 2022, supporting the importance of considering the age distribution of the excess deaths. Individuals in excess mortality in 2021 were younger than those in 2020, but the balance between the lower number of deaths of individuals and their greater remaining life expectancies globally yielded a greater number of YLL. The same rationale explains that despite a slightly lower number of excess deaths observed in 2023 as compared to the three previous years, the number of YLL remains high because the mean age of excess deaths was the lowest of the pandemic period, and the individuals in excess mortality had a mean remaining life expectancy of more than 13 years. Over the course of the pandemic, people aged less than 60 years old accounted for a growing part of the total burden, starting from less than 20% in 2020 up to 50% in 2023, attesting for a major impact of the pandemic not only on older people, but also on persons from 20 to 60 years old, particularly males.

The burden of the pandemic increased in France from 2020 to 2022, when considering both the occurrences of excess deaths and corresponding ages. Despite a decline in 2023, the global burden of excess mortality remained substantial and affected younger people than the previous years, mostly in the male population, yielding a high number of YLL. The occurrence of four consecutive years of excess mortality reported in this study builds an unprecedented picture: indeed, after a major episode of excess mortality, one might expect a drop in the following year(s), as was observed for instance in France, when considering the 15,000 excess deaths related to the 2003 heat wave, followed by a 23,000 drop of deaths in 2004 [28].

### Implications

Our study sheds light on the impact of the pandemic on mortality in France between 2020 and 2023. Investigating the causes of the increase over time evidenced in the study is of primary importance. Indeed, the co-occurrence in 2022 of critical features such as the achieved deployment of vaccination should have likely favoured a decreased burden as compared to earlier pandemic sub-periods. Actually, the mortality directly attributed to COVID-19 decreased over time during the pandemic period, with deaths officially attributed to the COVID-19 of 69,000, in 2020 [29], 60,895 in 2021 [30], 38,310 in 2022, and 5,572 in 2023 [31].

Opposed time trends between excess mortality estimated in the present study and the real dynamics of the COVID-19 threat (including the progressive immunity of the population protection after exposure and vaccination) suggest that indirect consequences of the pandemic, such as long-term effects on the health care system organization, may have contributed to increase mortality in 2022 and 2023. Numerous studies documented various critical pandemic-related changes in terms of health services: significant drops in hospitalizations for other causes than COVID-19 [32,33], delay or suspension of chronic disease management [34], delayed cancer diagnosis [35] likely leading to decreases of the long-term survival in these vulnerable sub-populations [36]. Other indirect contributions of the pandemic period may be thought. For example, the earliest period of the pandemic was associated with a massive decrease in the circulation of seasonal respiratory viruses such as influenza [37], whereas in 2022, France experienced a particularly high burden with two waves of influenza, one in March-April and one in December [38]. Different factors might have contributed to an indirect pandemic-related increased mortality over time of the pandemic period, and eventually yielded the maximum number of excess deaths estimated in 2022. Nevertheless, the persistence of excess mortality in 2023, and the burden on males under 60 years old remains unexplained and raises important concerns.

In 2020 and 2021 direct COVID-19 mortality was above excess mortality. The virus killed not only people who would not have died in the absence of a pandemic (amount of excess deaths) but also people who would have died from other causes, resulting in a decrease in other causes of death in 2020 and 2021 [29,30]. Unfortunately causes of death are not yet available in France for years 2022 and 2023, but the higher estimated number of excess deaths than that of COVID-19-attributed deaths likely reflects a long-term indirect impact of the pandemic, possibly increasing mortality related to modifications of patient management and healthcare system organization. A similar pattern for year 2022 was also reported in Germany [39] and Korea [40], suggesting a similar impact of the pandemic on healthcare systems and mortality not only in France. Further research is needed to better understand the various features contributing to this perturbation of mortality trends.

### Strengths and limitations

This study is the first to date to provide a complete and detailed toll of the COVID-19 pandemic on mortality in France from 2020 to 2023. A major strength of the study is the exhaustivity and the accuracy of the toll, precisely estimated by taking into account major parameters influencing mortality such as age- and sex-specific evolution over time and changes in population structure. As a nationwide cohort study, the present study is fully exhaustive for France, and the method used can be extended to other countries, provided comparable data details on mortality and population structure.

Some limitations must nevertheless be noted. The study predictions are based on the assumption that in the absence of the COVID-19 pandemic, expected mortality in the corresponding years would have followed the trends that were observed for the previous ten years, (including potential specific patterns for each age and sex) and this cannot be ascertained. Predicting four successive years inherently introduces a pattern of increasing uncertainty: the greater the time between prediction years and training period, the greater the uncertainty. Nevertheless, taking ten years of reference periods better considers regular trends than taking only five [41] or three years [7].

Estimates of YLL were calculated by applying the life expectancy of each person in excess mortality without considering underlying health conditions. The health status of the deceased was not documented at all in the database, but in average, their health condition was likely poorer than the average of the same-age and -sex population, with a resulting lower life expectancy. Indeed, many studies reported that deaths from COVID-19 were positively associated with the presence of comorbidities [42,43] and the total number of years of life lost was found to be 12% lower than expected in Hungary when comorbidities were taken into account [16]. In a previous study, Quast et al. decided to reduce the expected life expectancy by 25% to take this point into account [44]. The present study did not adopt any correction, so the reader must be cautious when interpreting the numeric YLL presented here, which may be overestimated. Nevertheless, the main study result on this topic concerns the increasing trend of progression over time, and at least the shape of this trend remains fully reliable. Estimates of the number of deaths officially attributed to COVID-19 should also be treated with caution. Indeed, some crucial parameters, such as the availability of tests and coding practices, varied greatly over the course of the pandemic, making strict interpretation of the official count uncertain. However, even if they are potentially underestimated, the 43,882 COVID-19 attributed deaths in 2022 and 2023 [31] are far from explaining the 70,484 excess deaths estimated for these two years.

### Conclusion and perspectives

This study provides the first comprehensive analysis of the impact of the pandemic on mortality in France between 2020 and 2023. Despite a fall in excess mortality in 2021 and 2023, the number of years of life lost remains at a critical level, reflecting substantial losses among individuals under 60 years old, particularly males. This four-year period of unprecedented excess mortality highly suggests a lasting indirect impact of the COVID-19 pandemic on healthcare system, even in 2023. Further research is essential to understand the reported trends and mitigate future risks.

## Data Availability

All data used in the study are available in open access from the French National Institute for Statistic and Economic Studies (Insee)
https://www.data.gouv.fr/fr/datasets/fichier-des-personnes-decedees/
https://www.insee.fr/fr/statistiques/series/103088458
https://www.insee.fr/fr/statistiques/6686513?sommaire=6686521

## Acknowledgements

The COVID-HOSP working group: Tristan Delory, Centre Hospitalier Annecy Genevois, Annecy, France; Fanny Duchaine, IRDES, Paris, France; Maude Espagnacq, IRDES, Paris, France; Gilles Hejblum, INSERM, Paris, France; Myriam Khlat, INED, Aubervilliers, France; Nathanaël Lapidus, INSERM, Paris, France; Sophie Le Cœur, INED, Aubervilliers, France; Elhadji Leye, INSERM, Paris, France; Paul Moulaire, INSERM, Paris FRANCE; Jonas Poucineau, INED, Aubervilliers, France.

## Author Contributions

GH and NL initiated and supervised the study; GH, NL, and PM designed the experimental plan; PM managed data and performed the analyses; GH, NL, and PM can take responsibility for the integrity of the data and the accuracy of the data analysis, PM is the guarantor; PM prepared the first draft of the manuscript; GH, NL, and PM contributed to interpretation of the data, critically revised the manuscript, and approved the final version.

## Ethics approval

This is an observational study using open data; no ethical approval is required

## Patient and public involvement

Patients and/or the public were not involved in the design, or conduct, or reporting, or dissemination plans of this research.

## References

1. World Health Organization. WHO COVID-19 dashboard. https://data.who.int/dashboards/covid19/deaths?n=c. Accessed 12 Jul. 2024.

2. Islam N. “Excess deaths” is the best metric for tracking the pandemic. BMJ 2022;376:o285.

3. Vanella P, Basellini U, Lange B. Assessing excess mortality in times of pandemics based on principal component analysis of weekly mortality data—the case of COVID-19. Genus 2021;77:16.

4. Kowall B, Stang A. Estimates of excess mortality during the COVID-19 pandemic strongly depend on subjective methodological choices. Herz 2023;48:180–83.

5. Nepomuceno MR, Klimkin I, Jdanov DA, Alustiza-Galarza A, Shkolnikov VM. Sensitivity Analysis of Excess Mortality due to the COVID-19 Pandemic. Popul Dev Rev 2022;48:279–302.

6. Ioannidis JPA, Zonta F, Levitt M. Flaws and uncertainties in pandemic global excess death calculations. Eur J Clin Invest n/a:e14008.

7. Levitt M, Zonta F, Ioannidis JPA. Comparison of pandemic excess mortality in 2020–2021 across different empirical calculations. Environ Res 2022;213:113754.

8. Wang H, Paulson KR, Pease SA, et al. Estimating excess mortality due to the COVID-19 pandemic: a systematic analysis of COVID-19-related mortality, 2020–21. The Lancet 2022;399:1513–36.

9. Karlinsky A, Kobak D. Tracking excess mortality across countries during the COVID-19 pandemic with the World Mortality Dataset. eLife 2021;10:e69336.

10. The Economist. Tracking covid-19 excess deaths across countries. 2021. https://www.economist.com/graphic-detail/coronavirus-excess-deaths-tracker. Accessed 12 Jul. 2024.

11. World Health Organization. Global excess deaths associated with COVID-19, January 2020 - December 2021. https://www.who.int/data/stories/global-excess-deaths-associated-with-covid-19-january-2020-december-2021. Accessed 12 Jul. 2024.

12. Blanpain N. 53,800 more deaths than expected in 2022: higher excess mortality than in 2020 and 2021. Insee Première. 2023; 1951. https://www.insee.fr/en/statistiques/7635866. Accessed 12 Jul. 2024.

13. European Mortality Monitoring Project. EuroMOMO (euromomo.eu) 2024. EUROMOMO. https://www.euromomo.eu/graphs-and-maps. Accessed 12 Jul. 2024.

14. Eurostat. Excess mortality statistics. https://ec.europa.eu/eurostat/databrowser/view/DEMO_MEXRTcustom_11946893/default/line?lang=en. Accessed 12 Jul. 2024.

15. Statement on the fifteenth meeting of the IHR (2005) Emergency Committee on the COVID-19 pandemic. https://www.who.int/news/item/05-05-2023-statement-on-the-fifteenth-meeting-of-the-international-health-regulations-(2005)-emergency-committee-regarding-the-coronavirus-disease-(covid-19)-pandemic. Accessed 28 Jun. 2024.

16. Ferenci T. Different approaches to quantify years of life lost from COVID-19. Eur J Epidemiol 2021;36:589–97.

17. Ugarte MP, Achilleos S, Quattrocchi A, et al. Premature mortality attributable to COVID-19: potential years of life lost in 17 countries around the world, January–August 2020. BMC Public Health 2022;22:54.

18. Pifarré I Arolas H, Acosta E, López-Casasnovas G, et al. Years of life lost to COVID-19 in 81 countries. Sci Rep 2021;11:3504.

19. Institut national de la statistique et des études économiques (French National Institute of Statistics and Economic Studies). Fichier des personnes décédées. https://www.data.gouv.fr/fr/datasets/fichier-des-personnes-decedees/. Accessed 12 Jul. 2024.

20. Institut national de la statistique et des études économiques (French National Institute of Statistics and Economic Studies). Séries Population et structure de la population | Insee. https://www.insee.fr/fr/statistiques/series/103088458. Accessed 12 Jul. 2024.

21. Institut national de la statistique et des études économiques (French National Institute of Statistics and Economic Studies). Espérances de vie - Bilan démographique 2022 | Insee. https://www.insee.fr/fr/statistiques/6686513?sommaire=6686521. Accessed 12 Jul. 2024.

22. Vandenbroucke JP, Elm E von, Altman DG, et al. Strengthening the Reporting of Observational Studies in Epidemiology (STROBE): Explanation and Elaboration. PLOS Med 2007;4:e297.

23. Heidari S, Babor TF, De Castro P, Tort S, Curno M. Sex and Gender Equity in Research: rationale for the SAGER guidelines and recommended use. Res Integr Peer Rev 2016;1:2.

24. Décret n°82-103 du 22 janvier 1982 relatif au répertoire national d’identification des personnes physiques. 1982.

25. Alicandro G, La Vecchia C, Islam N, Pizzato M. A comprehensive analysis of all-cause and cause-specific excess deaths in 30 countries during 2020. Eur J Epidemiol 2023;38:1153–64.

26. Fantin R, Barboza-Solís C, Hildesheim A, Herrero R. Excess mortality from COVID 19 in Costa Rica: a registry based study using Poisson regression. Lancet Reg Health – Am 2023;20. doi:10.1016/j.lana.2023.100451.

27. Barnard S, Chiavenna C, Fox S, et al. Methods for modelling excess mortality across England during the COVID-19 pandemic. Stat Methods Med Res 2022;31:1790–1802.

28. Toulemon L, Barbieri M. The mortality impact of the August 2003 heat wave in France: Investigating the ‘harvesting’ effect and other long-term consequences. Popul Stud 2008;62:39–53.

29. Naouri D. COVID-19: third leading cause of death in France in 2020, while other major causes of death decline. Études et Résultats. 2022 Dec;1250:1–7. https://drees.solidarites-sante.gouv.fr/publications-en-anglais/covid-19-third-leading-cause-death-france-2020-while-other-major-causes. Accessed 12 Jul. 2024.

30. Fouillet A, Ghosn W, Riviera C, Clanché F, Coudin E. Leading causes of death in France in 2021 and recent trends. Bull Épidémiol Hebd 202326554-69. https://beh.santepubliquefrance.fr/beh/2023/26/2023_26_1.html. Accessed 28 Jun. 2024.

31. COVID-19 Data Explorer. Our World Data. https://ourworldindata.org/explorers/coronavirus-data-explorer?zoomToSelection=true&time=2021-11-12.2024-01-17&facet=none&country=~FRA&pickerSort=asc&pickerMetric=location&Metric=Confirmed+deaths&Interval=Cumulative&Relative+to+Population=false&Color+by+test+positivity=false. Accessed 28 Jun. 2024.

32. Birkmeyer JD, Barnato A, Birkmeyer N, Bessler R, Skinner J. The Impact Of The COVID-19 Pandemic On Hospital Admissions In The United States. Health Aff Proj Hope 2020;39:2010–17.

33. Shah SA, Brophy S, Kennedy J, et al. Impact of first UK COVID-19 lockdown on hospital admissions: Interrupted time series study of 32 million people. eClinicalMedicine 2022;49:101462.

34. Smith M, Vaughan Sarrazin M, Wang X, et al. Risk from delayed or missed care and non-COVID-19 outcomes for older patients with chronic conditions during the pandemic. J Am Geriatr Soc 2022;70:1314–24.

35. Angelini M, Teglia F, Astolfi L, Casolari G, Boffetta P. Decrease of cancer diagnosis during COVID-19 pandemic: a systematic review and meta-analysis. Eur J Epidemiol 2023;38:31–38.

36. Maringe C, Spicer J, Morris M, et al. The impact of the COVID-19 pandemic on cancer deaths due to delays in diagnosis in England, UK: a national, population-based, modelling study. Lancet Oncol 2020;21:1023–34.

37. Rubin R. Influenza’s Unprecedented Low Profile During COVID-19 Pandemic Leaves Experts Wondering What This Flu Season Has in Store. JAMA 2021;326:899–900.

38. Santé Publique France (Public Health France). Bulletin épidémiologique grippe, semaine 18. Bilan préliminaire. Saison 2022-2023. https://www.santepubliquefrance.fr/maladies-et-traumatismes/maladies-et-infections-respiratoires/grippe/documents/bulletin-national/bulletin-epidemiologique-grippe-semaine-18.-bilan-preliminaire.-saison-2022-2023. Accessed 12 Jul. 2024.

39. Kuhbandner C, Reitzner M. Estimation of Excess Mortality in Germany During 2020-2022. Cureus 2023;15:e39371.

40. Han C, Jang H, Oh J. Excess mortality during the Coronavirus disease pandemic in Korea. BMC Public Health 2023;23:1698.

41. Msemburi W, Karlinsky A, Knutson V, Aleshin-Guendel S, Chatterji S, Wakefield J. The WHO estimates of excess mortality associated with the COVID-19 pandemic. Nature 2023;613:130–37.

42. Semenzato L, Botton J, Drouin J, et al. Chronic diseases, health conditions and risk of COVID-19-related hospitalization and in-hospital mortality during the first wave of the epidemic in France: a cohort study of 66 million people. Lancet Reg Health - Eur 2021;8:100158.

43. Working group for the surveillance and control of COVID-19 in Spain. The first wave of the COVID-19 pandemic in Spain: characterisation of cases and risk factors for severe outcomes, as at 27 April 2020. Eurosurveillance 2020;25:2001431.

44. Quast T, Andel R, Gregory S, Storch EA. Years of life lost associated with COVID-19 deaths in the United States. J Public Health 2020;42:717–22.

