## Supplementary for "Excess mortality and years of life lost from 2020 to 2023 in France: a cohort study of the overall impact of the COVID-19 pandemic on mortality"

### Supplementary Text 1.

#### Model selection

A Poisson regression model was used to estimate expected mortality in the pandemic period with 2010−2019 as reference period. Sex, age, and year were used as predictors to estimate mortality. Different interactions between these variables were tested, and AIC was used as the selection criterion to optimize goodness-of-fit and avoid overfitting (See supplementary Table 1). The simplest model tested expressed expected mortality as a function of age, sex and year:

$$\log\left(E\left[\text{Mortality}_{ijk}\right]\right) = \beta_0 + \beta_1 \cdot \text{Sex}_i + \beta_2 \cdot \text{Age}_j + \beta_3 \cdot \text{Year}_k$$

This model without interaction assumed similar effects of these variables across all different strata, and the corresponding AIC was 2,020,257. In a second model, the natural logarithm of the population was included as an offset value in order to take into account changes in the population structure, and yielded a decrease of the AIC to 332,359. In a third model, age was considered as factor to estimate coefficients for each age (categorical variable for age rounded to a year), therefore allowing a non-linear effect of age on mortality. With this third model AIC dropped to 79,182. Interaction between age and sex was added in a fourth model in order to enable a different effect of sex across age strata, and corresponding AIC decreased to 23,812. Finally, an interaction with year was added to estimate an annual trend in each stratum of age and sex, and the corresponding AIC dropped to 21,356. Therefore, this final model was selected and its analytical expression is the following:

$$\log\left(E\left[\text{Mortality}_{ijk}\right]\right) = \log\left(\text{Population}_{ijk}\right) + \beta_0 + \beta_1 \cdot \text{Sex}_i + \beta_2 \cdot \text{Age}_j + \beta_3 \cdot \text{Year}_k$$

$$+\beta_4 \cdot \text{Sex}_i \cdot \text{Age}_j + \beta_5 \cdot \text{Sex}_i \cdot \text{Year}_k + \beta_6 \cdot \text{Age}_j \cdot \text{Year}_k + \beta_7 \cdot \text{Sex}_i \cdot \text{Age}_j \cdot \text{Year}_k$$

The advantage of this model was to estimate age- and sex-specific expected numbers of deaths without any assumption about similarity of patterns across strata. This stratum-specific expected number of deaths allowed to estimate excess mortality and corresponding years of life lost.

**Supplementary Table 1.**

**Overview of the linear predictors used in the Poisson regression model with corresponding AIC and rationale, training period 2010–2019.**

| Linear predictor | AIC | Rationale |
|---|---|---|
| Year + Sex + $Age_c$ | **2,020,257** | Simplest model with year sex and age, average effect of each variable. |
| Year + Sex + $Age_c$ + log(Population) | **332,359** | Considers changes in population structure. |
| Year + Sex + $Age_f$ + log(Population) | **79,182** | Effect of age estimated for each level of age. |
| Year + Sex · $Age_f$ + log(Population) | **23,812** | Effect of sex estimated for each age. |
| Year · Sex · $Age_f$ + log(Population) | **21,356** | Annual trend estimated for each sex and age strata. |

$Age_c$ – age coded as a continuous variable; $Age_f$ – age coded as a factor (categorical variable)

**Supplementary Figure 1.**

**Study flow diagram.**

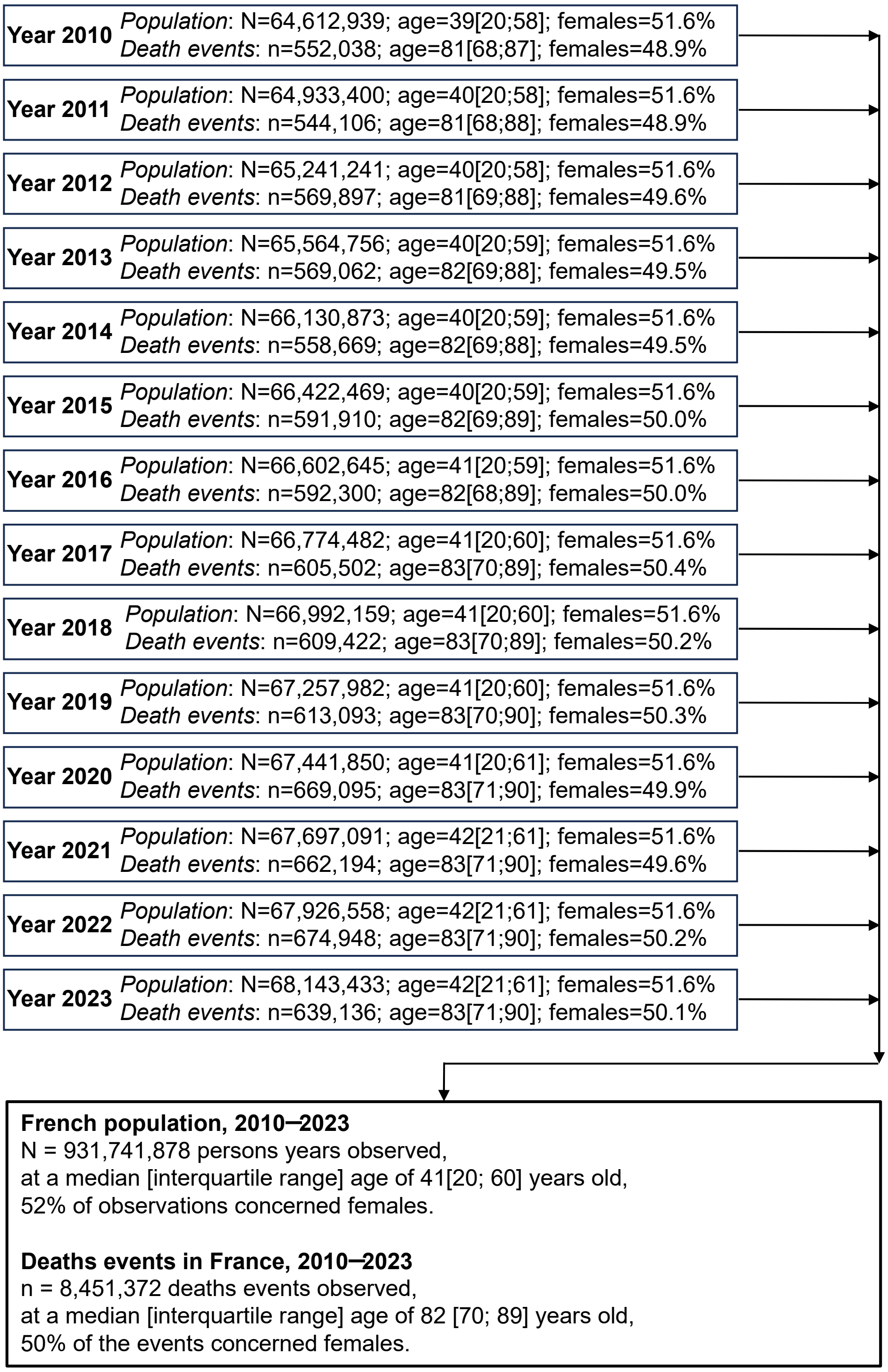


In each year-specific cell, N corresponds to the population size on January 1 of this given year. Age is reported by its median value [interquartile range], and the percentage of data concerning females is also reported. n corresponds to the total number of deaths events that occurred in France during this year. Over the study period (January 1, 2010 to December 31, 2023), 85,139 death events occurred abroad and were not included in this study on excess mortality in France.

**Supplementary Table 2.**

**Age of individuals in excess mortality and proportion of YLL under 60 years old, by sex, France, 2020−2023**

| Year | 2020 | 2021 | 2022 | 2023 |
|---|---|---|---|---|
| **Age of individuals in excess mortality*** | 83 [75; 89] | 78 [70; 84] | 81 [72; 89] | 76 [68; 82] |
| Males | 80 [73; 87] | 76 [68; 83] | 78 [70; 86] | 74 [65; 79] |
| Females | 86 [79; 91] | 80 [72; 85] | 84 [76; 91] | 78 [70; 84] |
| **Percentage of total YLL originating from persons in excess mortality aged less than 60 years old †** | 16.8% [14.7%; 18.8%] | 26.2% [24.2%; 28.0%] | 32.2% [30.5%; 34.0%] | 50.4% [46.6%; 54.3%] |
| Males | 22.9% [20.4%; 25.2%] | 34.5% [32.4%; 36.4%] | 41.0% [38.9%; 43.0%] | 60.5% [56.4%; 64.8%] |
| Females | 6.8% [2.9%; 10.6%] | 9.5% [5.2%; 13.6%] | 19.1% [15.9%; 22.3%] | 28.4% [19.0%; 36.8%] |

*Median age [interquartile range]
† Proportion [95% confidence interval]

**Supplementary Figure 2. Years of life lost by age and sex, France, 2020–2023.**

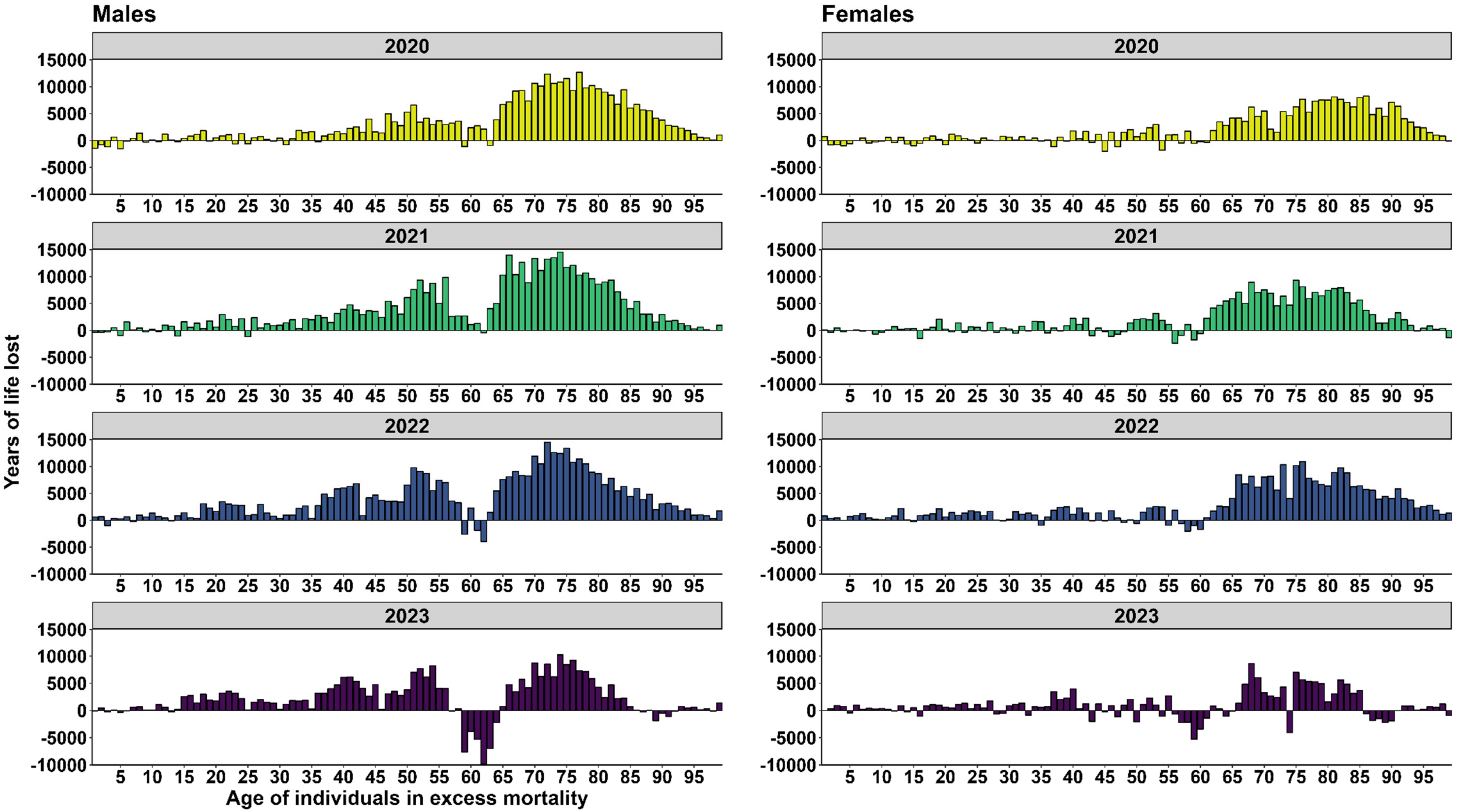


For each year between 2020 and 2023, histograms represent the number of years of life lost (y axis) by age of individuals in excess mortality concerned (x axis). The remaining life expectancy of people in excess mortality at each age had been summed to compare the contribution of each age. For instance, in 2023, males in excess mortality aged 40 years old contributed to about 5,000 years of life lost, close to those aged 71 years old, even if they were fewer in number, because their individual life expectancy was higher.
